# Cardiac Events Following JYNNEOS Vaccination for Prevention of Monkeypox

**DOI:** 10.1101/2022.11.13.22282258

**Authors:** Katie A Sharff, Thomas K Tandy, Paul F Lewis, Eric S Johnson

**Author notes:** Correspondence: Katie A. Sharff, Kaiser Permanente Northwest, Portland, Oregon.

## Abstract

**Purpose:** The JYNNEOS vaccine is an FDA-approved vaccine indicated for prevention of smallpox and monkeypox that is currently being distributed to high-risk individuals to combat the monkeypox public health emergency. The FDA safety data for JYNNEOS vaccine notes potential cardiac adverse events of special interest (AESI) following vaccination. The Advisory Committee on Immunization Practices (ACIP) stated that people with underlying heart disease or three or more major cardiac risk factors should be counseled about the theoretical risk for myopericarditis following vaccination with JYNNEOS. Our objective was to describe the incidence of cardiac AESIs in the Kaiser Permanente Northwest (KPNW) population who received a JYNNEOS vaccination for prevention of monkeypox.

**Methods:** We conducted a retrospective cohort study of 2,126 KPNW patients aged 12 or older who were vaccinated with at least 1 dose of JYNNEOS vaccine between July 14^th^ 2022 and October 10^th^ 2022. We searched KPNW databases, including the electronic health record (EHR), for diagnoses of elevated troponins, chest pain, myocarditis, pericarditis, myopericarditis, acute myocardial infarction, cardiac dysrhythmia, cardiac arrest, or ventricular fibrillation that were documented during any encounter (visit) using ICD-10-CM diagnosis codes. Two KPNW physicians reviewed all possible cases in the EHR to adjudicate cases as cardiac AESIs in alignment with the case definition used in the FDA safety data.

**Results:** There were 2,126 KPNW members who received a total of 3,235 JYNNEOS vaccine doses between July 14^th^ and October 10^th^. Encounter ICD10 diagnosis codes and troponin values identified 24 possible cardiac AESI cases. After physician adjudication, there were 10 confirmed cardiac AESI for an incidence of 3.1 per 1000 doses given with (exact 95% CI, 1.5 to 5.7). Of these 10 events, none could be attributed directly to the vaccination.

**Conclusion:** The incidence of cardiac adverse events following JYNNEOS vaccination for prevention of monkeypox is unknown and this is the first study to evaluate cardiac AESI following JYNNEOS vaccination in the 2022 monkeypox public health emergency. This retrospective cohort study of JYNNEOS vaccination for prevention of monkeypox identified 10 cardiac events that all had alternative explanations; and no hospitalizations or serious adverse outcomes were attributed to vaccination. This initial study provides timely information for the clinician counseling their patient with underlying cardiac risk factors on the low observed risk of cardiac events with JYNNEOS vaccination during the 2022 monkeypox public health emergency.

## Introduction

The US national monkeypox vaccine strategy was announced on June 28^th^ 2022. (1) The JYNNEOS vaccine is an FDA-approved vaccine indicated for prevention of smallpox and monkeypox that is currently being distributed to high-risk individuals to combat the monkeypox public health emergency via a subcutaneous (SC) or intradermal route (ID). (2) Patients initially received the SC injection in 0.5mL doses, however, health systems shifted to 0.1mL ID injections after it became apparent the current supply was not sufficient to address the demand.

The FDA safety data for JYNNEOS vaccine notes potential cardiac adverse events following vaccination. Clinical trials of a 2-dose series of the JYNNEOS vaccine administered subcutaneously included evaluation of cardiac adverse events of special interest (AESIs): any cardiac signs or symptoms, EKG changes determined to be clinically significant, or troponin-I elevated above 2 times the upper limit of normal. Cardiac AESIs occurred in 1.3% of JYNNEOS recipients and 0.2% of placebo recipients who were smallpox vaccine-naïve. (3) The Advisory Committee on Immunization Practices (ACIP) stated that people with underlying heart disease or three or more major cardiac risk factors should be counseled about the theoretical risk for myopericarditis following vaccination with JYNNEOS given the uncertain etiology of myopericarditis associated with replication-competent smallpox vaccines such as ACAM2000. (2)

Our objective was to describe the incidence of cardiac AESIs in the Kaiser Permanente Northwest (KPNW) population who received a JYNNEOS vaccination for prevention of monkeypox.

## Methods

We conducted a retrospective cohort study of 2,126 KPNW patients aged 12 or older who were vaccinated with at least 1 dose of JYNNEOS vaccine between July 14^th^ 2022 and October 10^th^ 2022. The patient must have been a KPNW member at the time of vaccination and through the 21-day follow-up period. Vaccination data were collected from internal KPNW records as well as Washington and Oregon state vaccination registries (e.g., if a KPNW member was vaccinated outside of a KPNW clinic).

We searched KPNW databases, including the electronic health record, for diagnoses of elevated troponins, chest pain, myocarditis, pericarditis, myopericarditis, acute myocardial infarction, cardiac dysrhythmia, cardiac arrest, or ventricular fibrillation that were documented during any encounter (visit) using ICD-10-CM diagnosis codes. The ICD-10-CM diagnosis codes included a comprehensive list developed by the latest version of Clinical Classification Software, categorized under “circulation” as follows: CIR005, CIR006, CIR009, CIR012, CIR017, CIR018. (4) Additionally, we searched for any encounter where a troponin test was reported based on procedure codes. All diagnoses and troponin tests had to occur within 21 days of JYNNEOS vaccination. Two KPNW physicians reviewed all possible cases to adjudicate cardiac AESIs based on the case definition used in the FDA safety data. (3) We calculated the incidence of cardiac adverse events of special interest per 1,000 vaccine doses administered using exact Poisson 95% confidence interval. KPNW’s institutional review board (IRB) approved the study.

## Results

KPNW members (n=2,126) received 3,235 JYNNEOS vaccine doses between July 14th and October 10th: 1033 received 1 dose (48.6%); 1,077 received 2 doses (50.7%); 16 patients received 3 doses (0.75%). Of the 2,126 members, 1,811 (85.2%) were male, 290 (13.6%) were female and 25 (1.2%) were non-binary or unknown. Ages ranged from 12 to 82 years.

Encounter ICD-10-CM diagnosis codes and troponin values identified 24 possible cardiac AESI events. Physician adjudication confirmed 10 cardiac AESI events for a positive predictive value of 41.7%. The incidence was 3.1 per 1,000 doses (exact 95% CI, 1.5 to 5.7). Of these 10 events, none could be attributed directly to the vaccination (i.e., no primary AESI events). We were unable to estimate the incidence of cardiac AESI by route of administration because the states’ registry data (external to KPNW) did not capture route. Of the 10 confirmed cardiac AESI events, 7 occurred in male patients, 2 occurred in female patients and 1 occurred in a non-binary patient. Of the patients who suffered a cardiac AESI event, 9 of the 10 (90%) had a history of at least one cardiac risk factor (Table). The most common presenting symptoms included chest pain (60%) and palpitations (50%). Seven patients (70%) were seen exclusively in the outpatient setting and three patients (30%) presented to the emergency department (ED). No patients were hospitalized because of the event. All cases were self-limited with no attributable deaths.

**Table.** Summary of cardiac adverse events of special interest (AESI) following JYNNEOS vaccination

| Age | Cardiac History | Dose | Route | Onset after Vaccination | Cardiac Symptoms | Evaluation Location | EKG | Diagnosis |
| --- | --- | --- | --- | --- | --- | --- | --- | --- |
| 30-39 | None | 1 | ID | 19 days | Chest pain<br>Palpitations | Office | Yes-Sinus | Anxiety |
| 20-29 | Tobacco use | 1 | SC | 12 days | Sustained<br>Chest pain x 3 days | UC | No | Pneumonia |
| 30-39 | Amphetamine use | 1 | SC | 5 days | Palpitations | ED | Yes-Sinus tachycardiac | Palpitations attributed to drug use |
| 30-39 | Obesity | 1 | SC | 13 days | Chest pain | ED | Yes-Sinus bradycardia | Post-operative chest wall pain |
| 30-39 | Obesity | 1 | SC | 14 days | Chest Pain | Video then Office | Yes- Ectopic atrial rhythm | Post-COVID19 chest pain, triggered by vaccine |
| 40-49 | HTN<br>Obesity<br>Angina<br>Hyperlipidemia | 1 | SC | 21 days | Intermittent chest pain | Video | Yes-Sinus | Angina |
| 50-59 | HTN<br>Tobacco Use | 2 | ID | 19 days | Chest pain | UC | Yes- sinus arrhythmia | Troponin WNL<br>Diagnosed with chest pain and discharged |
| 60-69 | Paroxysmal AFIB<br>Obesity<br>HTN<br>Atherosclerosis | 1 | ID | 4 days | Palpitations | Office | No | Paroxysmal AFIB, medication adjustment |
| 60-69 | Obesity<br>HTN<br>Atherosclerosis | 1 | ID | 7 days | Palpitations | Office to ED | Yes-Sinus tachycardia | Palpitations & poorly controlled Hypertension. BP medication prescribed |
| 60-69 | AFIB<br>Atherosclerosis | 2 | ID | 15 days | Palpitations<br>Weakness<br>SOB | Telephone | No | Paroxysmal AFIB |

ID=Intradermal; SC=Subcutaneous AFIB= Atrial fibrillation; HTN=Hypertension; SOB=Shortness of breath; UC=Urgent Care; ED=Emergency Department; WNL=Within normal limits; BP=Blood pressure

## Discussion

This is the first study to evaluate cardiac AESI following JYNNEOS vaccination in the 2022 monkeypox public health emergency. In this retrospective cohort study of 3,235 doses of JYNNEOS vaccine doses administered, we identified 10 cardiac AESI for an incidence of 3.1 per 1,000 doses administered.

The FDA safety data for JYNNEOS vaccine notes potential cardiac adverse events but the case definition of these events is broad. (3) ACIP reports they have not detected an increased risk of myopericarditis in recipients of JYNNEOS but recommends counseling patients with underlying heart disease about theoretical risk. (2)

Of the AESI events that we identified by ICD-10 or troponin values, only 41.7% met the FDA definition of cardiac AESI. When these events were adjudicated by physician chart review, we verified that none of the 10 cases were attributable to the JYNNEOS vaccination. Our clinical chart review highlights the importance of physician adjudication to understand the true attributable incidence of adverse events following vaccination. It is important to highlight that outpatient visits accounted for 70% of our cases which may be overlooked by other pharmacovigilance systems that look only at ED and hospital visits. (5)

This initial study provides timely information for the clinician counseling their patient with underlying cardiac risk factors on the low observed risk of cardiac events with JYNNEOS vaccination for prevention of monkeypox during the 2022 public health emergency. This retrospective cohort study of JYNNEOS vaccination for prevention of monkeypox identified 10 cardiac events that all had alternative explanations, and no hospitalizations or serious adverse outcomes were attributed to vaccination. Future research should evaluate incidence of cardiac AESI by dose number, route of administration and the contribution of pre-existing conditions to event rates.

## Data Availability

All data produced in the present study are available upon reasonable request to the authors

